# Droplet microfluidics-based detection of very rare antibiotic-resistant subpopulation in clinical isolates of *Escherichia coli* from bloodstream infections

**DOI:** 10.1101/2025.03.05.25323409

**Authors:** Sagar N. Agnihotri, Nikos Fatsis-Kavalopoulos, Jonas Windhager, Maria Tenje, Dan I. Andersson

## Abstract

Population heterogeneity in bacterial phenotypes, such as antibiotic resistance, is increasingly recognized as a medical concern. Heteroresistance (HR) occurs when a predominantly susceptible bacterial population harbors a rare resistant subpopulation. During antibiotic exposure, these resistant bacteria can be selected and lead to treatment failure. Standard antibiotic susceptibility testing (AST) methods often fail to reliably detect these subpopulations due to their low frequency, highlighting the need for new diagnostic approaches. Here, we present a droplet microfluidics method where bacteria are encapsulated in droplets containing growth medium and antibiotics. The growth of rare resistant cells is detected by observing droplet shrinkage under microscopy. We validated this method for three clinically important antibiotics in *Escherichia coli* isolates obtained from bloodstream infections and showed that it can detect resistant subpopulations as infrequent as 10^−6^ using only 200 to 300 droplets. Additionally, we designed a multiplex microfluidic chip to increase the throughput of the assay.

## Introduction

Antibiotic resistance presents a significant challenge in modern medicine^1^ where escalating resistance has increased the demand for more precise and faster Antibiotic Susceptibility Testing (AST)^2,3^. Although numerous technologies have emerged aiming to rapidly diagnose antibiotic resistance^4–10^, most of these rapid ASTs are inefficient in detecting resistant subpopulations present at very low frequencies (10^-6^ or 10^-7^) due to the sheer number of bacteria that need to be analyzed. Heteroresistance (HR) is one such phenotype characterized by a predominantly susceptible bacterial population harboring rare resistant subpopulations^11–13^. When antibiotics are administered, creating selective pressure, these resistant subpopulations can grow and potentially result in treatment failure^14–17^.

HR poses a particular challenge for traditional AST diagnostics, which typically assess the resistance characteristics of homogenous cell populations. In contrast, HR involves quantifying resistance in a rare subpopulation of cells, making it difficult to detect using standard diagnostic methods. Current clinical diagnostics such as disk diffusion, Etests, and broth microdilution often fail to identify HR^18–21^, underscoring a critical gap in our ability to manage and treat bacterial infections effectively. Missed HR can complicate treatment and have dire implications for patient health^22–25^. The only method currently used to detect HR is the Population Analysis Profile (PAP) test, which is time-consuming and labor-intensive and therefore rarely used. As a result, clinicians and researchers struggle to reliably and rapidly detect HR phenotypes^12,13,26^.

HR in *Escherichia coli* is of special interest as this species is both a WHO priority pathogen and the leading cause of sepsis and bacteremia worldwide^27–31^. *E. coli* bacteremia is a severe infection with high hospital mortality rates^27^ and any possible treatment failure due to HR could have detrimental effects on patients’ health. Addressing the challenge of accurately detecting HR phenotypes in *E. coli* clinical isolates was the primary goal of this study. The technology presented here utilizes droplet-based microfluidics to identify resistant subpopulations associated with clinical HR strains. Droplet-based microfluidics is frequently employed to encapsulate mammalian cells or bacteria for single-cell analysis^32,33^ and AST^34–40^. This technique can encapsulate cells in pL to fL-sized droplets by adjusting the cell concentration in the initial cell suspension^41^. In addition, these droplets can be manipulated^42,43^ and sorted^44^ in the microfluidic chip to achieve different permutations and combinations, similar to biological assays in a standard laboratory. There are very few studies using droplet-based microfluidics to detect heteroresistance^45,46^, these studies employ single-cell encapsulation per droplet resulting in a lot of empty droplets, and to detect low subpopulation frequency of 10^-6^, these methods will require around 5 to 6 million droplets along with transfer in between chips. Hence, there is a need for a microfluidic method that will reduce the number of droplets required along with no in-between chip transfer steps. To improve the detection limit with just 200 to 300 droplets and have detection on the same chip, we encapsulate hundreds to thousands of bacteria per droplet in the presence of antibiotics. By tracking size changes in hundreds of such droplets, we can identify rare subpopulations that continue to grow even at antibiotic concentrations inhibiting most cells. This growth is quantified through osmotically induced size changes in the droplets, driven by the metabolic activity of the growing cells^47,48^.

We validated this novel approach on multiple clinical *E. coli* isolates, demonstrating its effectiveness in detecting HR compared to the reference PAP test method. The result is a scalable and multiplexable microfluidic chip that can reliably identify HR across three classes of antibiotics commonly used in clinical settings at clinically relevant frequencies and within clinically relevant timeframes. This new technology promises to improve our ability to identify infections caused by HR bacteria and it paves the way for implementation of advanced HR diagnostics in the healthcare system.

## Results

### Overview of the technology and analysis

We used droplet-based microfluidics to detect the resistant sub-population in clinical HR strains against single and multiple antibiotics. Previous work showed that droplets with encapsulated bacterial cells that grow and are in contact with droplets that do not contain growing cells (here referred to as “inert” droplets) shrink in size due to osmosis^47,48^. We used this feature to detect the resistant subpopulation at breakpoint antibiotics concentration (i.e. the concentration used to define whether an infection by a particular bacterial strain/isolate is likely to be treatable in a patient) in clinical isolates that showed a HR phenotype as determined by PAP tests. Between 100 to 3000 bacterial cells were encapsulated in each droplet depending on the antibiotics used, droplet size, and the frequency of the resistant subpopulation to detect. The droplets containing the resistant subpopulation that continued to grow in the presence of antibiotics showed a 5-10% reduction in droplet size compared to the droplets without any growth (Fig. 1), allowing detection of the resistant subpopulation down to a frequency of 10^-6^ by analyzing only 350 droplets over 24 hours. To detect HR against a single antibiotic, we used a standard T-junction set-up with a single inlet and a single incubation chamber (Fig. 1a). This design was then scaled to detect multiple antibiotics by using multiple droplet generators and multiple incubation chambers, forming a multiplex chip (Fig. 1b). Each incubation chamber had a capacity of 2000 to 3500 droplets in the range of 180 to 220 µm diameter.

**Figure 1:**
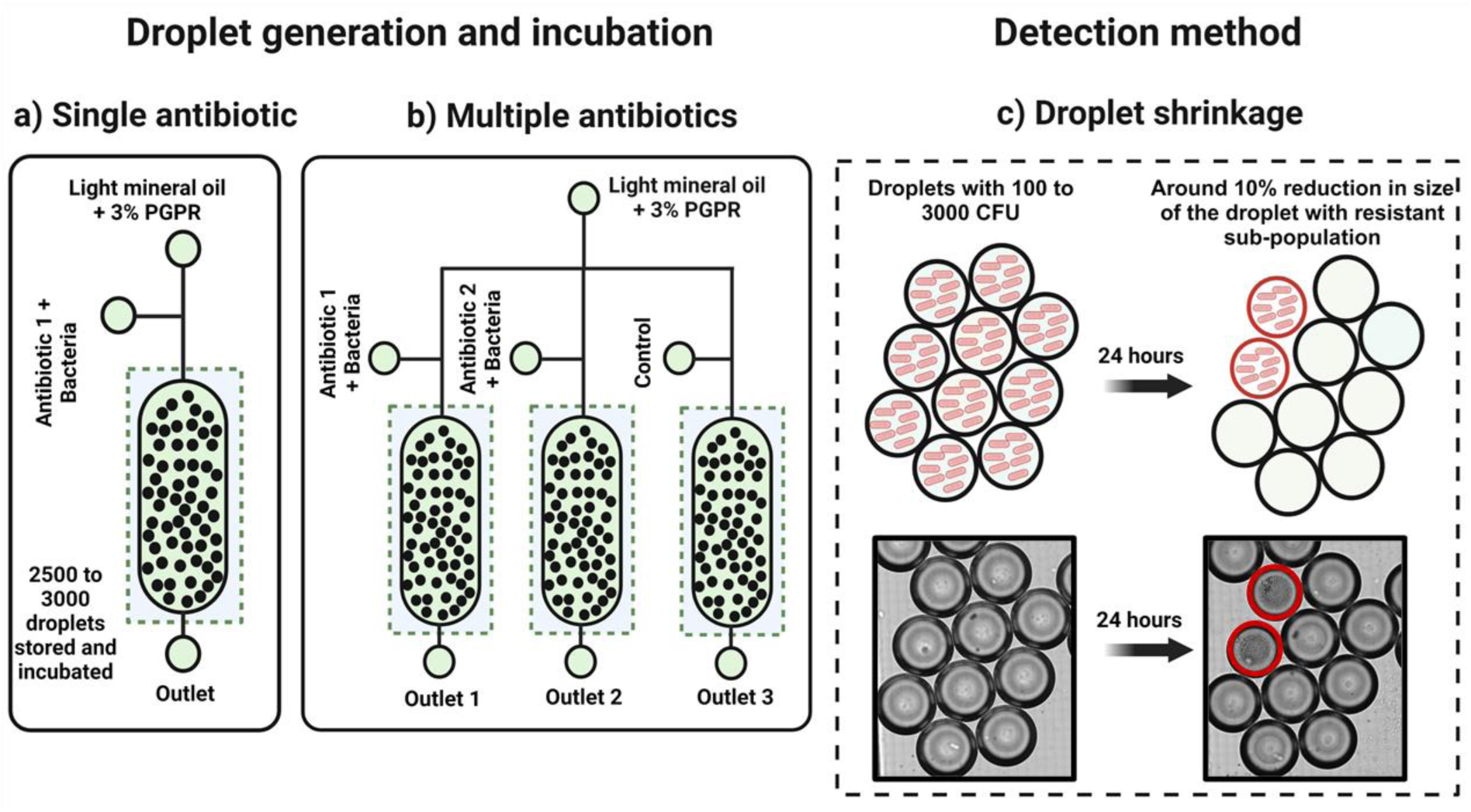
Schematic showing an overview of the technology. For detection against a single antibiotic, a standard T-junction droplet generator is used with one droplet incubation chamber (Fig. 1a) while against multiple antibiotics several droplet generators and incubation chambers are used (Fig. 1b). Each droplet contains antibiotics and around 400 to 3000 encapsulated bacteria. Droplets with a resistant subpopulation of bacteria that can grow in the presence of antibiotics show an approximately 10% reduction in size which is detectable by microscopy (Fig. 1c).

### Size differences between empty and bacteria-filled droplets

To evaluate how the technology performs for the detection of bacterial growth, we first optimized the ratio of filled to empty droplets by varying the concentration of bacteria in the initial droplet-forming suspension. We used three different CFU/mL concentrations of the *E. coli* laboratory strain MG1655 encapsulated in MH broth without antibiotics, generating different encapsulation ratios: 10^4^ = approximately 1% bacteria encapsulated and 99% empty droplets, 10^5^ = approximately 26% bacteria encapsulated and 74% empty droplets, and 5×10^5^ = approximately 69% bacteria encapsulated and 31% empty droplets. As shown in Fig. 2 (a-c) droplet shrinkage could be detected with all tested ratios of filled-to-empty droplets and the number of droplets displaying shrinkage compared well with the expected number of cell-containing droplets, as calculated at the encapsulation stage. Strikingly, even when only 1% of the droplets contained *E. coli*, this growth could be detected. The microscopic images at 0 and 24 hours for each set of experiments are shown in fig. 2a (ii and iii) - 2c (ii and iii) indicating empty droplets (yellow-bordered circle) and bacteria-containing droplets (red-bordered circle).

**Figure 2:**
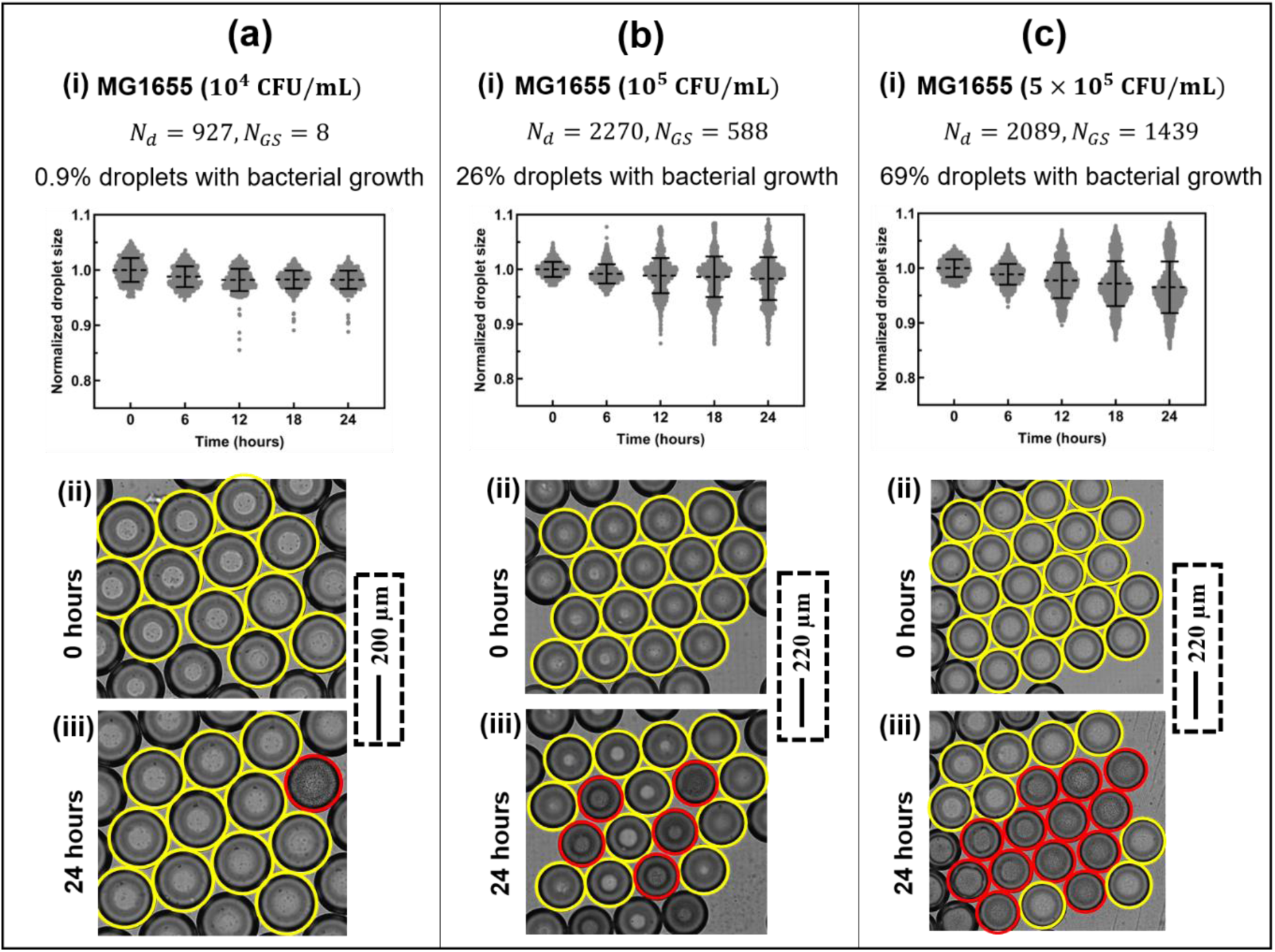
MG1655 in MH broth with different colony forming units (CFU) per mL to have varying numbers of droplets with bacterial encapsulation and growth. **(a)** (i) 10^4^ CFU/mL (ii) Image at 0 hours (iii) Image at 24 hours along with the dimensions. **(b)** (i) 10^5^ CFU/mL (ii) Image at 0 hours (iii) Image at 24 hours **(c)** (i) 5×10^5^ CFU/mL (ii) Image at 0 hours (iii) Image at 24 hours. Droplets with yellow-colored boundaries represent the droplets without growth while droplets with red-colored boundaries represent droplets showing bacterial growth.

### Effect of antibiotics on growth and droplet size with susceptible MG1655

To ensure that the droplet-microfluidic platform could correctly replicate such results and did not report false negatives, we performed a control experiment where susceptible *E. coli* (MG1655) was exposed to either cefotaxime (CTX) or gentamicin (GEN) at 2 mg/L and 0.5 mg/L, respectively. No growth was observed with either CTX or GEN (Fig. 3 a-b). These plots demonstrate that there is no growth and associated shrinkage in any of the droplets when susceptible bacteria are exposed to high antibiotic levels (above the minimum inhibitory concentration, MIC).

**Figure 3:**
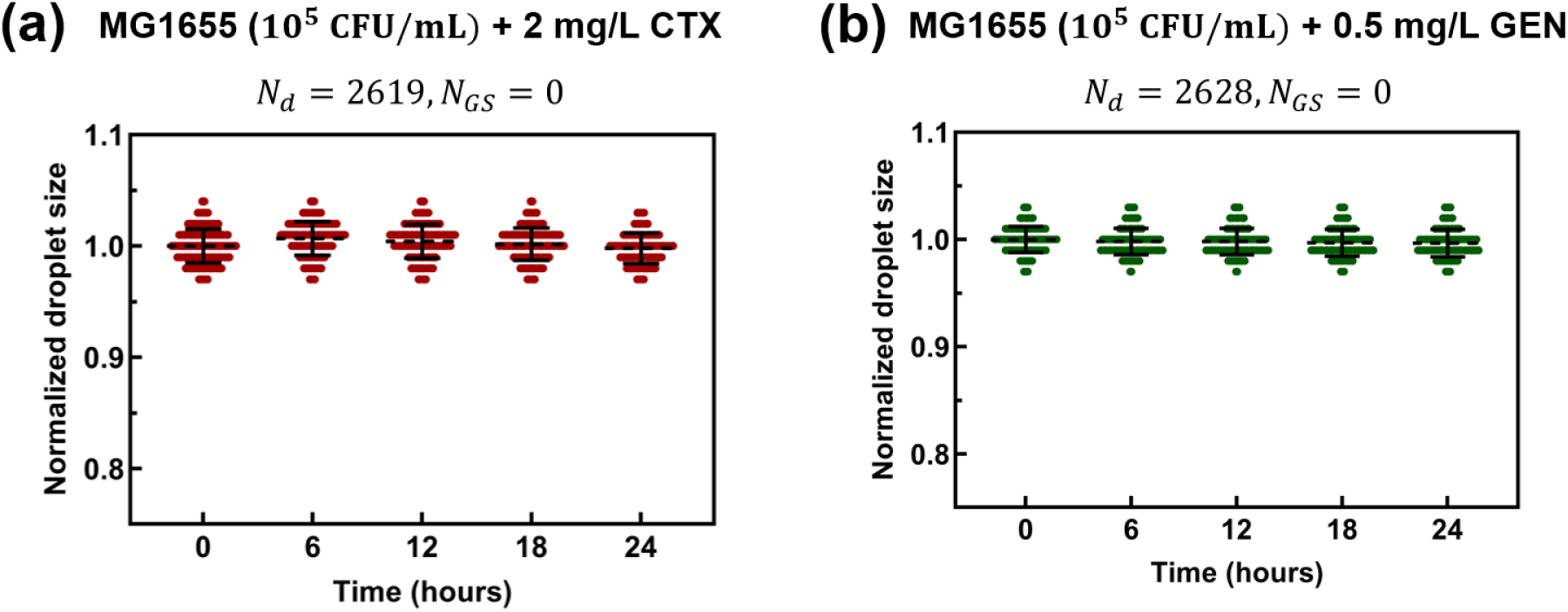
Effect of two different antibiotics on susceptible MG1655 with 10^5^ CFU/mL. **(a)** 2 mg/L CTX **(b)** 0.5 mg/L GEN. N_d_ represents the number of droplets imaged and analyzed and N_GS_ represents droplets that show bacterial growth and shrinkage of the droplet.

### Size differences between droplets containing susceptible and resistant *E. coli*

In the previous experiments, we confirmed that a difference in size could be observed between droplets that contain growing bacteria and those that do not. Next, we aimed to explore if a resistant subpopulation could be detected when co-encapsulated with a main population of susceptible cells. To this end, we spiked suspensions of susceptible MG1655 with resistant *E. coli* at different frequencies (10^-3^, 10^-5^, and 10^-6^) and encapsulated the cells in droplets comprising MH broth with antibiotics. Three different antibiotic classes were used, namely β-lactams (cefotaxime, CTX at 2 mg/L), aminoglycosides (gentamicin, GEN at 4 mg/L), and tetracyclines (tetracycline, TET at 5 mg/L) where the concentrations used were above the clinical breakpoint for the respective antibiotic to ensure that only droplets containing resistant cells would shrink.

As shown in Fig. 4a-g and Table 1, the number of droplets imaged and analyzed (N_d_) and number of droplets with bacterial growth and associated shrinkage (N_GS_) varied between 1519 to 2611 and 3 to 204, respectively. Based on determinations of CFUs/mL for the susceptible and resistant populations before mixing, we knew the expected frequencies of resistant bacteria encapsulated. To calculate the frequency of resistant bacteria detected in the experiment we used the following formula: number of droplets with shrinkage / (total number of droplets × average number of bacteria per droplet). This allowed us to assess how well the experiments determined the frequency of the resistant subpopulation by comparing the spiked-up frequency to that experimentally observed. Table 1 summarizes these numbers, and as can be seen the frequencies are in good agreement with each other. We note that the experimentally calculated frequency slightly overestimates the spiked frequency, primarily due to one reason. Thus, despite statistically calculating the average number of bacteria per droplet, variation can arise due to bacterial sedimentation within the tubing connecting the syringe pump to the microfluidic chip. Taken together, these experiments show that we can correctly detect the resistant subpopulations even when their frequency is as low as 10^-6^ (Fig. 4).

**Figure 4:**
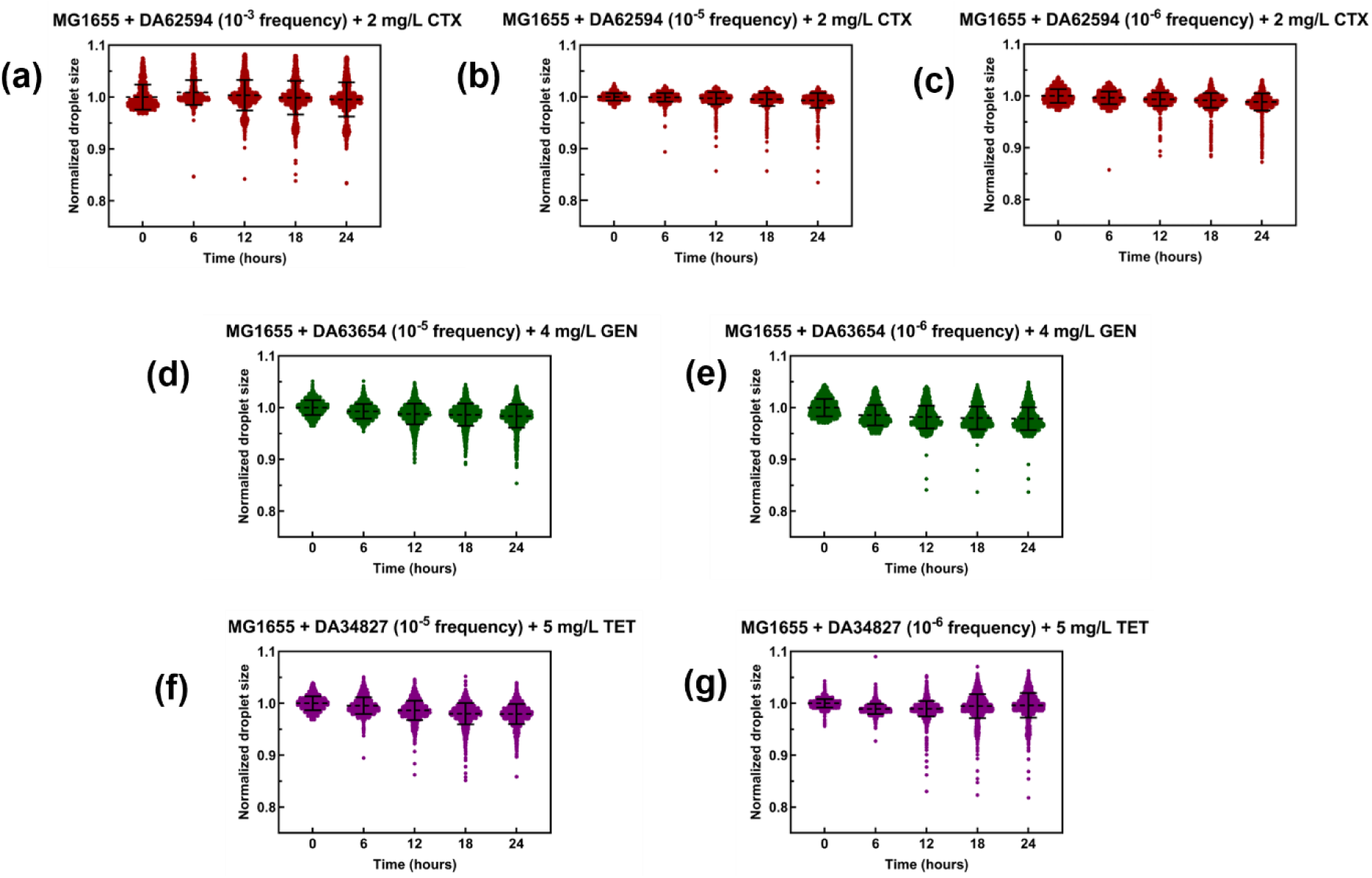
Spiking susceptible *E. coli* with resistant *E. coli* in the presence of antibiotics to detect the subpopulation of resistant *E. coli*. For the different cases, normalized droplet sizes are plotted against time. **(a)** MG1655+ CTX and resistant strain DA62594 with a 10^-3^ frequency in the presence of 2 mg/L CTX. CFU/mL = 2.5×10^7^. **(b)** MG1655+CTX resistant strain DA62594 with a 10^-5^ frequency in the presence of 2 mg/L CTX. CFU/mL= 5×10^8^. **(c)** MG1655+CTX resistant strain DA62594 with a 10^-6^ frequency in the presence of 2 mg/L CTX. CFU/mL = 5×10^8^. **(d)** MG1655+GEN resistant strain DA63654 with a 10^-5^ frequency in the presence of 4 mg/L GEN. CFU/mL= 5×10^8^. **(e)** MG1655+GEN resistant strain DA63654 with a 10^-6^ frequency in the presence of 4 mg/L GEN. CFU/mL= 5×10^8^. **(f)** MG1655+TET resistant strain DA34827 with a 10^-5^ frequency in the presence of 5 mg/L TET. CFU/mL= 5×10^8^. **(g)** MG1655+TET resistant strain DA62594 with a 10^-6^ frequency in the presence of 5 mg/L TET. CFU/mL= 5×10^8^.

**Table 1:** Data showing the number of droplets analyzed (N_d_), droplets with shrinkage (N_GS_), average, and total volume of droplets analyzed in the case of susceptible MG1655 with different frequencies of resistant *E. coli* subjected to CTX, GEN, and TET respectively. Spiked-up frequencies and experimentally calculated frequencies are compared.

| Droplet content | $N_d$ | $N_{GS}$ | Bacteria encapsulated per droplet | Spiked frequency | Experimentally calculated frequency |
| --- | --- | --- | --- | --- | --- |
| MG1655 + DA62594+ 2mg/L CTX | 1519 | 204 | 65 | $10^{-3}$ | $2.0 \times 10^{-3}$ |
| MG1655 + DA62594+ 2mg/L CTX | 1649 | 60 | 1630 | $10^{-5}$ | $2.2 \times 10^{-5}$ |
| MG1655 + DA62594+ 2mg/L CTX | 2611 | 37 | 2350 | $10^{-6}$ | $6.0 \times 10^{-6}$ |
| MG1655 + DA63654+ 4mg/L GEN | 2534 | 123 | 2000 | $10^{-5}$ | $2.4 \times 10^{-5}$ |
| MG1655 + DA63654+ 4mg/L GEN | 1757 | 3 | 2320 | $10^{-6}$ | $7.4 \times 10^{-7}$ |
| MG1655 + DA34827+ 5mg/L TET | 2077 | 170 | 2290 | $10^{-5}$ | $3.5 \times 10^{-5}$ |
| MG1655 + DA34827+ 5mg/L TET | 2520 | 80 | 2065 | $10^{-6}$ | $1.5 \times 10^{-5}$ |

### Detection of resistant subpopulations in HR clinical isolates of *E. coli*

Finally, we wanted to assess if the method could reproduce the previous sensitivities to detect HR for clinical isolates. Here, we used three different strains that were also tested by PAP. In this set of experiments, we used the two antibiotics CTX and GEN at 2 mg/L and 0.5 mg/L, respectively (schematically shown in Fig. 5a.) Two strains of *E. coli* (DA63082 and DA63744) displaying HR to CTX, and two *E. coli* strains (DA63082 and DA63340) showing HR to GEN, as determined by PAP tests were used and each experiment was performed two times (N_1_ and N_2_), with the data plotted side by side in Fig. 5 (b-e). As summarized in Fig. 5 a-e and Table 2, the number of droplets imaged and analyzed (N_d_) and the number of droplets with bacterial growth and associated shrinkage (N_GS_) varied between 2101 to 3176 and 38 to 140, respectively. We compared the frequencies of HR determined by the PAP tests to the values from the experiments, and as can be seen in Table 2 there was some minor variation, but overall, there was good agreement.

**Figure 5:**
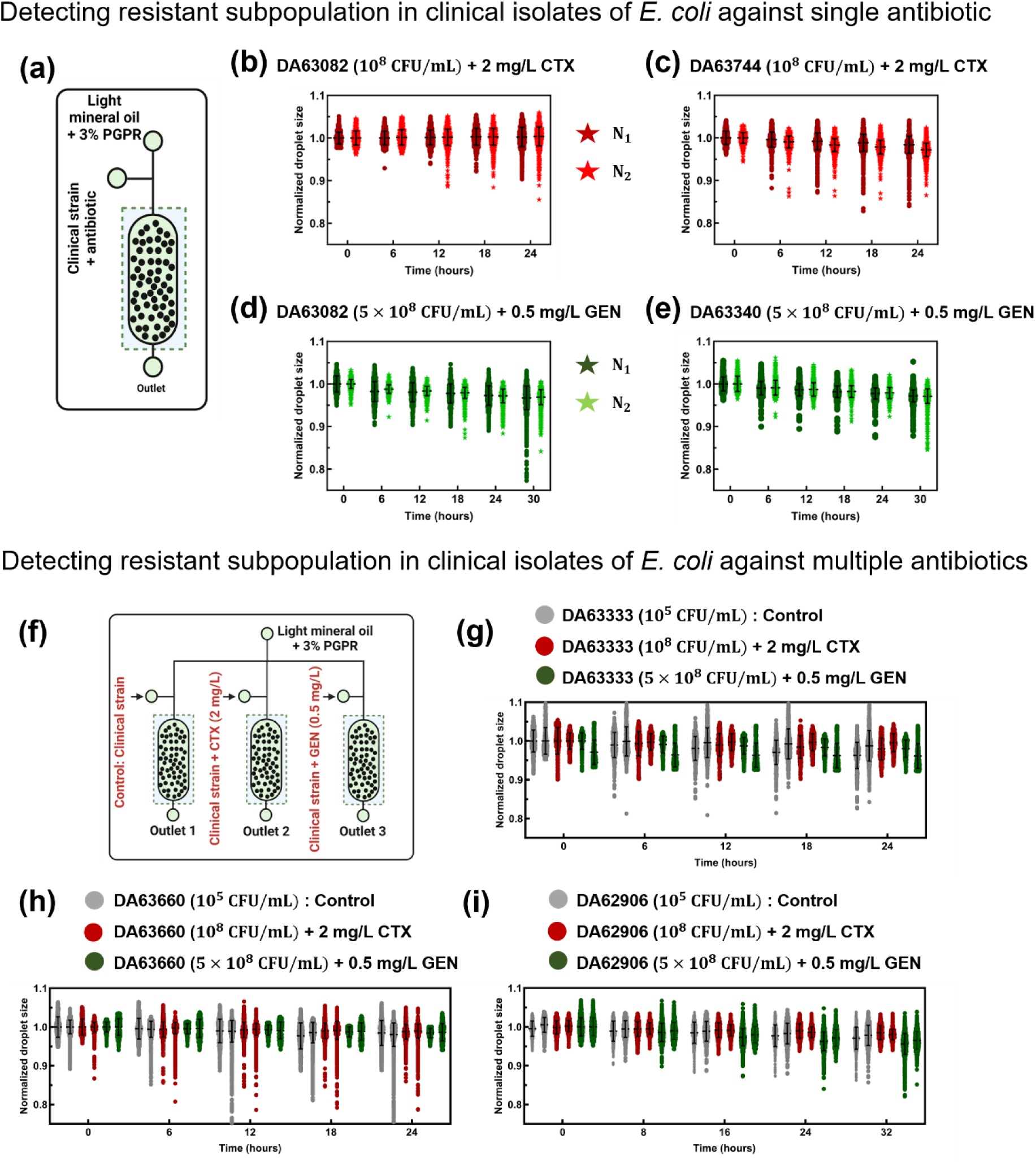
Detection of resistant subpopulations using clinical isolates of *E. coli* with HR against a single antibiotic (a-e). (a) Schematic of the single chip with inlets and outlets. **(b)** DA63082 HR against CTX. **(c)** DA63744 HR against CTX. **(d)** DA63082 HR against GEN. **(e)** DA63340 HR against GEN. Detection of resistant subpopulations using clinical isolates of *E. coli* against multiple antibiotics using the multiplex chip (f-i). (f) Schematic of the multiplex chip with an inlet of control, CTX, and GEN. **(g)** DA63333 is non-HR against both CTX and GEN. DA63333 is tested using a multiplex chip, the normalized droplet size is plotted against time and only the control experiment showed shrinkage. At the same time, there was no shrinkage and bacterial growth observed in the CTX and GEN droplets. **(h)** DA63660 shows HR against CTX and non-HR against GEN. DA63660 is tested using a multiplex chip. The normalized droplet size is plotted against time and both the control and CTX experiments showed shrinkage while there was no shrinkage and bacterial growth observed in the GEN droplets. **(i)** DA62906 shows HR against GEN and non-HR against CTX. DA62906 is tested using a multiplex chip. The normalized droplet size is plotted against time and both the control and GEN experiments showed shrinkage while there was no shrinkage and bacterial growth observed in the CTX droplets. Each strain was tested in two independent experiments (N_1_ and N_2_) with data plotted side by side demonstrating droplet size reduction caused by the bacterial growth.

**Table 2:** Data showing the number of droplets analyzed (N_d_), droplets with shrinkage (N_GS_), and bacteria encapsulated per droplet in the case of clinical isolates of *E. coli* showing HR against CTX and GEN in case of single and multiplex chip. PAP test frequencies are compared to the experimentally calculated values.

| Droplet content | | $N_d$ | $N_{GS}$ | Bacteria encapsulated per droplet | PAP test | Experimentally calculated frequency |
| --- | --- | --- | --- | --- | --- | --- |
| **Detecting resistant subpopulation HR clinical isolates in *E. coli* against single antibiotics** |  |  |  |  |  |  |
| DA63082 + 2mg/L CTX | $N_1$ | 2614 | 38 | 469 | $9.4 \times 10^{-6}$ | $3.1 \times 10^{-5}$ |
| | $N_2$ | 2982 | 67 | 468 | | $4.8 \times 10^{-5}$ |
| DA63744 + 2mg/L CTX | $N_1$ | 3058 | 46 | 458 | $2.2 \times 10^{-6}$ | $3.3 \times 10^{-5}$ |
| | $N_2$ | 3637 | 71 | 434 | | $4.5 \times 10^{-5}$ |
| DA63082 + 0.5 mg/L GEN | $N_1$ | 2767 | 82 | 2297 | $1.0 \times 10^{-5}$ | $1.3 \times 10^{-5}$ |
| | $N_2$ | 2101 | 57 | 1051 | | $2.6 \times 10^{-5}$ |
| DA63340 + 0.5 mg/L GEN | $N_1$ | 2377 | 123 | 1622 | $2.0 \times 10^{-5}$ | $3.2 \times 10^{-5}$ |
| | $N_2$ | 2172 | 140 | 1631 | | $4.0 \times 10^{-5}$ |

| Detecting resistant subpopulation HR clinical isolates in *E. coli* against multiple antibiotics |  |  |  |  |  |  |
| --- | --- | --- | --- | --- | --- | --- |
| Non-HR strain DA63333 |  |  |  |  |  |  |
| DA63333 (control) | N<sub>1</sub> | 523 | 43 | 0.37 | N/A | N/A |
|  | N<sub>2</sub> | 1121 | 53 | 0.32 |  | N/A |
| DA63333 + 2mg/L CTX | N<sub>1</sub> | 607 | 0 | 182 | < 10<sup>-7</sup> | 0 |
|  | N<sub>2</sub> | 1493 | 0 | 147 |  | 0 |
| DA63333 + 0.5 mg/L GEN | N<sub>1</sub> | 515 | 0 | 985 | < 10<sup>-7</sup> | 0 |
|  | N<sub>2</sub> | 1679 | 0 | 735 |  | 0 |
| CTX HR strain DA63660 |  |  |  |  |  |  |
| DA63660 (Control) | N<sub>1</sub> | 2549 | 217 | 0.36 | N/A | N/A |
|  | N<sub>2</sub> | 9788 | 170 | 0.08 |  | N/A |
| DA63660 + 2mg/L CTX | N<sub>1</sub> | 5260 | 59 | 197 | 2.6×10<sup>-5</sup> | 5.6×10<sup>-5</sup> |
|  | N<sub>2</sub> | 7705 | 53 | 121 |  | 5.7×10<sup>-5</sup> |
| DA63660 + 0.5 mg/L GEN | N<sub>1</sub> | 573 | 0 | 2260 | < 10<sup>-7</sup> | 0 |
|  | N<sub>2</sub> | 1076 | 0 | 2335 |  | 0 |
| GEN HR strain DA62906 |  |  |  |  |  |  |
| DA62906 (Control) | N<sub>1</sub> | 1712 | 98 | 0.46 | N/A | N/A |
|  | N<sub>2</sub> | 1590 | 57 | 0.44 |  | N/A |
| DA62906 + 2mg/L CTX | N<sub>1</sub> | 1797 | 0 | 483 | < 10<sup>-7</sup> | 0 |
|  | N<sub>2</sub> | 2114 | 0 | 481 |  | 0 |
| DA62906 + 0.5 mg/L GEN | N<sub>1</sub> | 2451 | 83 | 2080 | 2.0×10<sup>-5</sup> | 1.6×10<sup>-5</sup> |
|  | N<sub>2</sub> | 2788 | 58 | 2065 |  | 1.0×10<sup>-5</sup> |

### Multiplex chip to detect clinical HR against multiple antibiotics

To analyze the same clinical strains simultaneously against multiple antibiotics, we developed a multiplex chip comprised of a joint oil inlet and three independent aqueous phase inlets to generate and store three different kinds of droplets. For this set of experiments, we encapsulated (i) the clinical *E. coli* strain to be analyzed at 10^8^ CFU/mL in 2 mg/L CTX, (ii) 5×10^8^ CFU/mL of the clinical *E. coli* strain in 0.5 mg/L GEN and (iii) 10^5^ CFU/mL in MH broth (control) to test if the strains were HR against CTX or GEN or non-HR as shown schematically in Fig. 5f. All these experiments were performed two times (N_1_ and N_2_) and plotted side by side in Fig. 5 (g-i). The experiments are described in more detail in the following subsections.

### Detecting non-HR strain DA63333

In the first set of experiments, we used a clinically isolated non-HR strain DA63333 in the multiplex chip. The number of droplets in all the experiments varied from 523 to 1679 as shown in Fig. 5g and Table 2. Only the control experiments showed shrinkage of ∼40-50 of the analyzed droplets. There was no growth and shrinkage observed in the case of CTX and GEN droplets, similarly to what was observed in the PAP test, demonstrating that the DA63333 strain does not display HR against CTX and GEN.

### Detecting CTX HR strain DA63660

In the second set of experiments, we used the clinical CTX HR strain DA63660 in the multiplex chip following the same procedure. Here, the number of droplets in all the experiments varied from 573 to 9788 as shown in Table 2 and the control experiments showed shrinkage of 217 and 170 droplets respectively in N_1_ and N_2_. The two experiments involving DA63660 encapsulated with 2 mg/L CTX resulted in shrinkage of 59 and 53 droplets, as shown in Figure 5h. Using the number of droplets showing shrinkage, we calculated the experimentally observed frequency of resistant bacteria and compared it with the PAP test results. Both assays were in good agreement in demonstrating CTX HR in this strain. There was no growth and shrinkage observed in the case of DA63660 encapsulated in 0.5 mg/L GEN as plotted in Figure 5h, which is similar to the result from the PAP test.

### Detecting GEN HR strain DA62906

In the third set of experiments, we used the clinical GEN HR strain DA62906 in the multiplex chip. The number of droplets in all the experiments varied from 1590 to 2788 as shown in Table 2. The control experiments showed shrinkage of 98 and 57 droplets respectively in N_1_ and N_2_. There was no growth and shrinkage observed in the case of DA62906 encapsulated in 2 mg/L CTX as plotted in Figure 5i, which is similar to the PAP test. The two experiments involving DA62906 encapsulated in 0.5 mg/L GEN resulted in shrinkage of 83 and 58 droplets as plotted in Figure 5i. From the number of droplets showing shrinkage, we calculated the experimentally observed frequency of resistant cells and compared it with PAP test results. Also in this case there was good agreement between the assays in demonstrating GEN HR in this *E. coli* strain. Taken together, our multiplex chip can distinguish between non-HR, CTX HR, and GEN HR strains, and the multiplex chip can easily be further developed to increase analysis throughput by including more inlets for additional antibiotics and bacterial strains.

## Discussion

The global emergence of antibiotic resistance has reduced our options and ability to efficiently treat bacterial infections. Heteroresistance poses an especially significant challenge because of the difficulties associated with detecting the presence of a resistant subpopulation within a predominantly susceptible bacterial population using current AST methods ^11,12^. Furthermore, the gold standard PAP method for detection of HR is rarely used in clinical microbiology laboratories due to high costs, difficulty in automation and a slow turnaround time of around 2 days, which is too slow for blood infections^30^.

A recent study showed that HR in *E. coli* isolates from bloodstream infections ranged in frequencies from 0.4% to 42.7%, depending on the antibiotic, and that HR bacteria were generally misdiagnosed as susceptible by disk diffusion. Most importantly, for piperacillin-tazobactam (TZP) and gentamicin (GEN) the risk of worsened outcome (i.e., increased mortality and transfer to ICU) was strongly increased for infections caused by HR *E. coli* ^49^. These findings stress the importance of developing new automated methods with improved ability to rapidly detect rare resistant subpopulations.

To address this need, we developed an assay utilizing droplet-based microfluidics. This method involves encapsulating approximately 100 to 3,000 bacteria per droplet, depending on the type of antibiotic used, droplet size, and the frequency of the resistant subpopulation. When the bacterial population inside the droplet is exposed to antibiotics the resistant subpopulation grows, resulting in a 5-10% shrinkage of the droplet compared to droplets with only susceptible bacteria where growth is inhibited. The shrinkage of droplet size is a reliable indicator of the growth of resistant bacteria and can be precisely quantified by microscopy.

Our experiments demonstrate that droplet shrinkage (indicating bacterial growth) and swelling (indicating no growth) depend on the number of inert droplets in the incubation chamber (Fig. 2). When only a few droplets contain growing bacteria, they are mainly in contact with droplets without bacterial growth resulting in maximum shrinkage due to surrounding droplets supporting mass transfer. In contrast, when most droplets contain growing bacteria, the size difference is less noticeable as there are fewer surrounding droplets for the mass transfer process. This sensitivity at lower frequencies is a particular strength for HR detection.

Our method outpaces the current PAP test in terms of detection time and automation capabilities. Results can be obtained within 12 to 24 hours (depending on the antibiotic), which compared to the PAP test takes at least one additional day. This approach holds promise for identifying HR bacteria and subsequently adjusting treatment to reduce complications, failure, and death^50^. This method also significantly improves the detection limit for HR as compared to single encapsulation droplet-based standard AST methods since to detect lower frequencies, such as 10^−6^ or 10^-7^, they would require approximately 5.5×10⁶ and 5.5×10^7^ droplets, respectively. In contrast, our method detects subpopulations at a frequency of 10^−6^ using only 200 to 300 droplets, representing a 20,000-fold reduction in the number of droplets needed. Additionally, we have incorporated multiplexing into this microfluidic chip, allowing the detection of resistant subpopulations across multiple antibiotics simultaneously, making it a more efficient approach. Beyond its time-saving benefits, our method offers complete automation potential via integration with a flow control system and microscope. It can also facilitate the isolation of rare antibiotic-resistant phenotypes for further genotypic and phenotypic analysis when combined with active sorting methods like acoustic and electrical sorting based on droplet size disparity ^51^. Additionally, unlike the standard PAP test, which requires approximately 25 mL of MH broth media and antibiotics per agar plate, our approach requires just 5 µL, resulting in a 1/5000 reduction in the consumption of antibiotics. This work examined three clinically important antibiotics used for *E. coli* bloodstream infections, and future work will be aimed at applying this method to other bacterial species and clinically used antibiotics for which HR has been observed^11–13^.

## Methods and Material

### Microfluidic device design, fabrication, and setup

The microfluidic device consists of two parts: the droplet generation part is a standard T-junction with an inlet of oil (light mineral oil, Sigma-Aldrich with 3% PGPR (polyglycerol polyricinoleate) as a surfactant, Danisco) having a channel width of 160 µm and an inlet of aqueous phase (*E. coli* in MH broth + antibiotics) with a 100 µm wide channel. The height of all channels is 160 µm. The droplet generation section generates droplets in the range of 180 to 220 µm in diameter. The droplet storage and incubation section can hold around 2000 to 3500 droplets at the present capacity of the device. In the case of the multiplex chip, similar dimensions are used along with multiple droplet generators and incubation chambers. The microfluidic device mold was 3D printed using an ASIGA printer. The master mold was then used for PDMS (Polydimethylsiloxane) casting with a 10:1 ratio of base to curing agent from Sylgard 184. Then the PDMS replica was punched at inlets and outlets with a 1 mm punch (Welltec) and bonded to a glass slide (epredia) with a plasma oven (Diener Electronic, Germany). Syringe pumps (Cetoni, Nemesys pumps, Germany) were used to drive all fluids and the whole microfluidic setup was kept inside the stage-top incubator (OKOlab) maintained at 37°C and 80% humidity for the experiments.

### Bacterial strains and culturing conditions

Mueller-Hinton (Difco) medium was used for growth in broth and agar plates. Antibiotics (Gentamicin, Cefotaxime, and Tetracycline) were purchased from Sigma-Aldrich. For every experiment, bacteria frozen in 10% DMSO stocks were streaked on Mueller Hinton (MH) agar plates and incubated at 37°C overnight. Single colonies from the platers were inoculated in 1 mL MH broth, incubated at 37°C, 199 rpm agitation, overnight, and subsequently used for each experiment. Antibiotic stocks were prepared fresh from powder stock (Sigma Aldrich) in sterile PBS (Sigma-Aldrich) and used within 16 h of preparation.

When the clinical isolates were incubated in the droplet system in the presence of 4mg/L GEN, they exhibited filamentous growth. This growth pattern interfered with the system’s capacity to identify empty droplets. Therefore, we adjusted the protocol to use a lower concentration of 0.5 mg/L for this specific combination, which allowed for effective segmentation of droplets containing growth.

### Microscopy

We used an Olympus inverted microscope (IX73, Olympus) with a 10x objective, a motorized stage for x-y-z direction, and a camera (C11440, HAMAMTSU) for imaging of the droplets over 24 hours. The microfluidic device was placed in a stage-top incubator (OKOlab) with controlled temperature and humidity. Once droplets in the incubation chamber reached a stable state, each location was marked with a motorized stage in the x-y direction. Each location was then imaged every hour or every two hours (in case of multiplex chip) for the next 24 hours or 30 hours (in case of GEN HR strains).

### Image processing pipeline

The images that were obtained every hour were processed using a custom computational pipeline implemented in Python (see Code Availability) to determine the shrinkage in droplet diameter due to bacterial growth. Briefly, raw grayscale images were smoothened (Gaussian 31×31 kernel, standard deviation: 5), opened (21×21 kernel), min-max-normalized, and converted to unsigned 8-bit integer format. Droplet boundaries were identified by thresholding pre-processed images using Otsu’s method (three classes: “droplet boundary”, “other droplet content”, “background”). The binary “droplet boundaries” masks were subjected to a circle Hough transform, droplet centroids were located by identifying peaks in accumulator-normalized Hough space, and maximum droplet radii were computed using a Euclidean distance transform of the “droplet boundaries” masks. Preliminary binary droplet masks were generated by labeling both “droplet boundaries” and “droplet disks” (given by droplet centroids and maximum radii) as regions corresponding to droplets. Droplets were then segmented based on a Euclidean distance transform of the preliminary droplet masks using a droplet centroid-seeded Watershed algorithm. Droplets touching the border of an image were excluded. Droplet label masks were exported as TIFF files, and tabular droplet information (accumulator values, centroids, radii) was saved in CSV format. For downstream analysis, droplet sizes were normalized by the average droplet diameter at 0 hours and plotted against time every 6 hours for 24 to 30 hours.

## Data Availability

All data produced in the present study are available upon reasonable request to the authors

## Author Contribution

Conceptualization: SNA, NFK, MT, DIA. Technology design and fabrication: SNA. Formal analysis: SNA, NFK, Funding acquisition: MT, DIA. Investigation: SNA, NFK. Methodology: SNA, NFK, MT, DIA. Writing – original draft: SNA. Writing – review and editing: SNA, NFK, JW, MT, DIA. Image processing pipeline (conceptualization & implementation): JW.

## Acknowledgments

This research was funded by grants from the Swedish Research Council (grant 2021-02091) and Knut and Alice Wallenberg Foundation (grant 2018.0168) to DIA and the European Research Council (ERC) under the European Union’s Horizon Europe research and innovation programme (PHOENIX grant agreement No 101043985) to MT. We also acknowledge help from the BioImage Informatics Facility, a unit of the National Bioinformatics Infrastructure Sweden NBIS, with funding from SciLifeLab, National Microscopy Infrastructure NMI (VR-RFI 2019-00217), and Chan Zuckerberg Initiative DAF (DAF2021-225261, DOI 10.37921/644085ggkbos, an advised fund of Silicon Valley Community Foundation, DOI 10.13039/100014989). U-PRINT carried out 3D printing: Uppsala University’s 3D printing facility in the scientific field of medicine and pharmacy. We acknowledge Myfab Uppsala for providing facilities and experimental support. Myfab is funded by the Swedish Research Council (2019-00207) as a national research infrastructure.

## Supplementary information

### Effect of droplet size on *E. coli* growth

To determine how *E. coli* grows in different sizes of droplets, we used the procedure as shown in Fig. S2. Droplets of different sizes (a total of 15 different sizes of droplets in the range of 20 µm to 200 µm in diameter) were generated using standard flow-focusing devices as shown in SI figure 1a. Light mineral oil with 3% PGPR as a surfactant was used as a continuous phase and MG1655 in MH broth media is used as a dispersed phase to generate droplets continuously. For droplet generation, the initial colony forming unit (CFU) per mL of MG1655 in MH broth was calculated for different sizes of the droplets generated to have a single/double encapsulation of MG1655 cells per droplet. These generated droplets were then transferred to a total of 6 different Eppendorf tubes for different incubation times (0, 2, 3, 4, 8, and 24 hours) as shown in SI figure 1b. After the incubation time in each case, protocols as shown in SI figure 1c were performed. Finally, the resulting media was put on an agar plate for 24 hours at 37°C to determine the final CFU.

**Figure S1:**
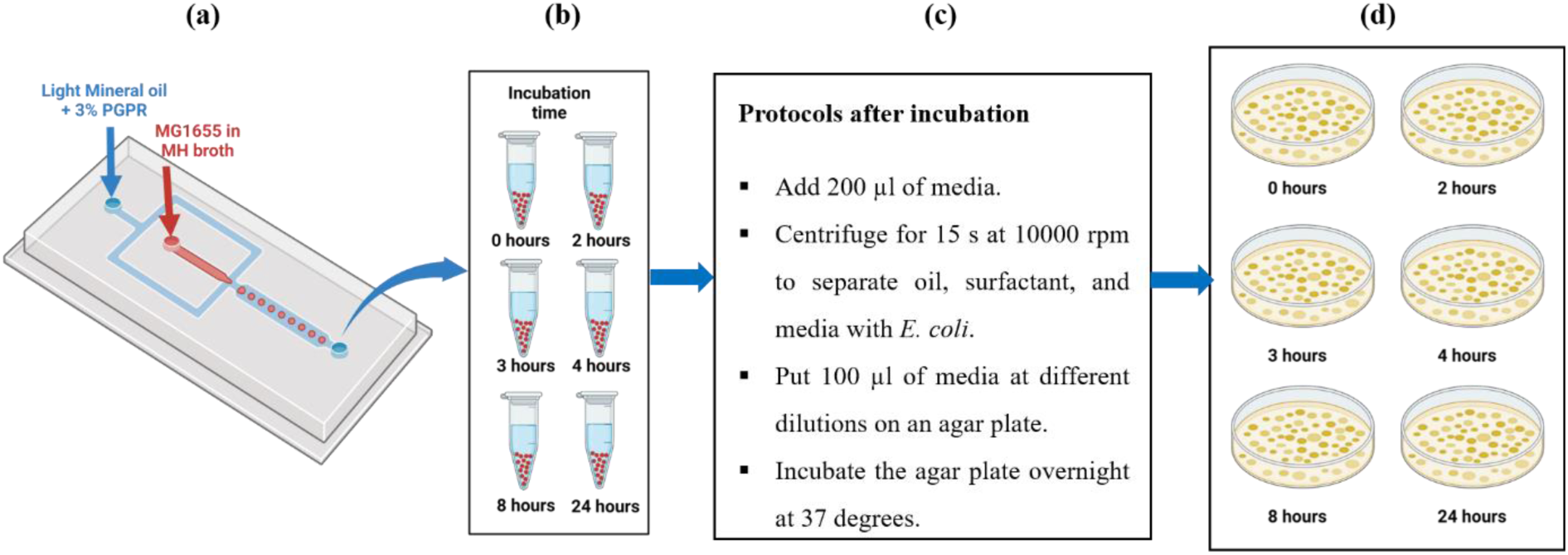
Schematic showing steps implemented to find out the effect of droplet size on MG1655 growth. **(a)** Schematic showing flow-focusing device used for droplet generation, where light mineral oil with 3% PGPR is used as a continuous phase and MG1655 in MH broth is used as a dispersed phase. **(b)** Droplets generated are transferred to Eppendorf tubes for incubation. **(c)** Protocols to be implemented after incubation time. **(d)** The resulting CFU is determined by growing MG1655 on agar plates incubated at 37°C for 24 hours.

After 24 hours on agar plates, colonies of MG1655 are counted and plotted in SI Fig. 2a. To normalize the growth, we divided CFU at all the time points by CFU at 0 hours as shown in SI Fig. 2b. It shows that in droplet sizes below 130 µm, MG1655 growth becomes stagnant after 8 hours, while in the case of droplet sizes above 140 µm, MG1655 growth is still in the exponential phase. Taken together, we conclude that for having substantial growth of MG1655 over a longer period, it is best to use a droplet diameter over 140 µm. Hence, for all our experiments, we have used droplet diameter in the range of 180 to 220 µm.

**Figure S2:**
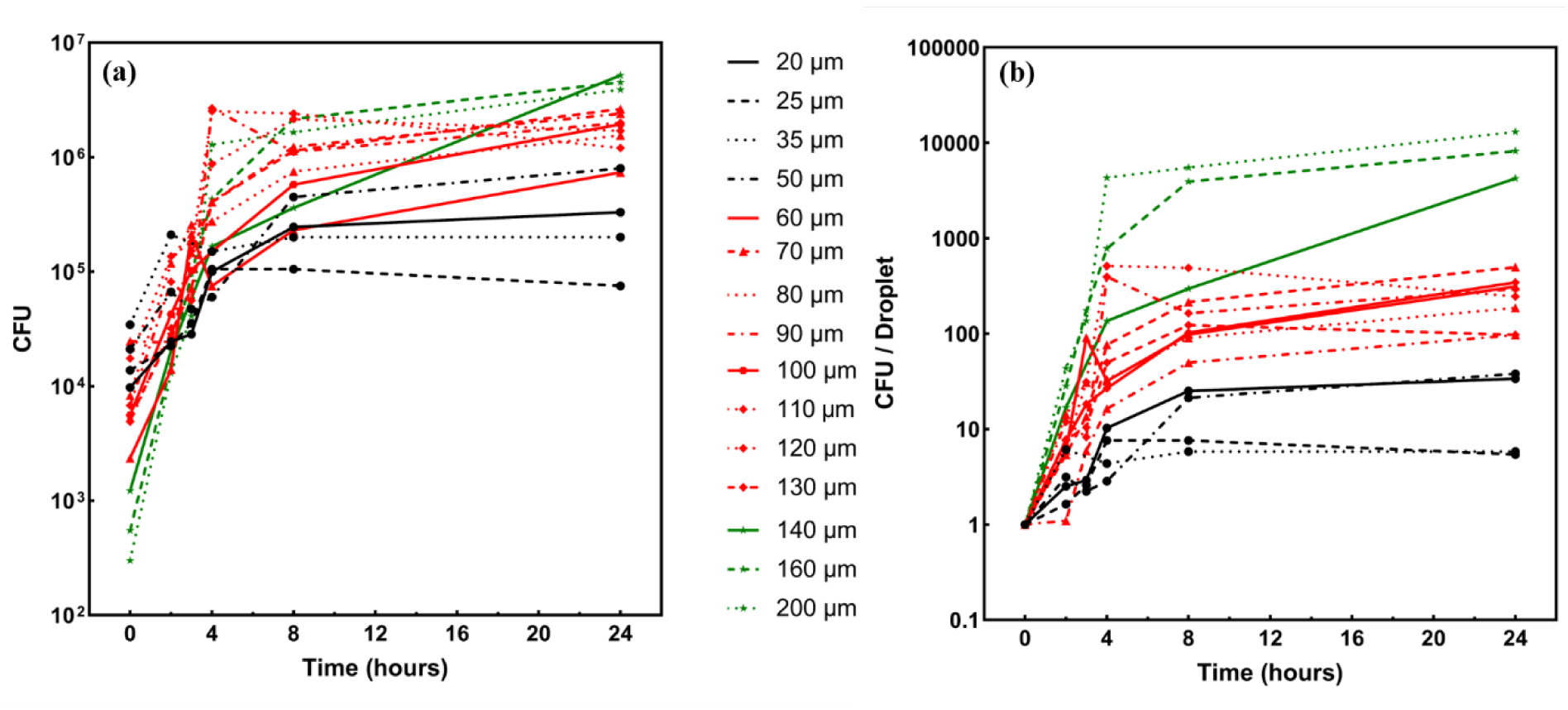
Effect of droplet size on MG1655 growth. **(a)** CFU of MG1655 plotted against time for various droplet diameters. **(b)** Normalized CFU of MG1655 plotted against time for various droplet diameters.

### Fabrication of flow-focusing devices

We used a flow-focusing device to generate droplets of various sizes. The flow-focusing devices of various sizes were fabricated using standard soft lithography. Firstly, a Cr/glass mask was fabricated using a mask writer, and a total of 15 designs were printed on the mask. Following that, a Si wafer was cleaned using a plasma oven, and SU8 2050 was spin-coated to a thickness of 80 µm followed by standard pre-baking and post-baking protocols. The pattern was transferred to the SU8 with a mask aligner and the non-exposed SU8 was developed with a standard SU8 developer. The developed SU8 mold was then hard-baked at 120°C. The PDMS Sylgard 184 along with the curing agent in the ratio of 10:1 was molded on the fabricated SU8 master and cured in an oven at 65°C. Once cured, inlets and outlets were punched and the PDMS devices were bonded to a glass slide using a plasma oven.

### Calculation for comparing experimentally calculated heteroresistance (HR) frequency and PAP test frequency

Encapsulation of *E. coli* or any cells in droplets is governed by Poisson distribution^1^ and it depends on the initial concentration of cells (here CFU/mL) and size of the droplet. In our case, we wanted to encapsulate 2000 or more CFU per droplet in the case of Gentamicin (GEN) and Tetracyclin (TET). We have therefore used 5×10^8^ CFU/mL when detecting resistant frequencies of 10^-5^ and 10^-6^. In the case of analyzing HR for Cefotaxime (CTX), we wanted to encapsulate 400 or more CFU per droplet and therefore used 10^8^ CFU/mL for detecting resistant frequencies of 10^-5^ and 10^-6^. When detecting resistant cells at 10^-3^ frequency we used only 2.5×10^7^ CFU/mL at the encapsulation stage.

The detailed calculations are as follows: When a 5×10^8^ CFU/mL of *E. coli* in MH broth is compartmentalized in a droplet of 4.7 nL size, the average number of CFU per droplet (λ) can be calculated as:

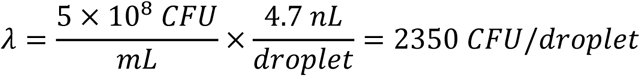

The same formula is used to calculate the average CFU per droplet in all cases. The λ obtained like this is used to calculate the experimentally observed HR frequency as follows:

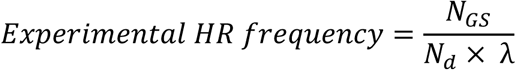

N_GS_ is the number of droplets showing bacterial growth and shrinkage, while N_d_ is the number of droplets analyzed.

